# A Longitudinal Neuromelanin MRI Processing Framework Reduces Measurement Variability and Improves Precision in Parkinson’s Disease

**DOI:** 10.64898/2026.07.24.26358895

**Authors:** Victoria Madge, Vladimir S. Fonov, David Araujo, Lydia Chougar, Dumitru Fetco, Madeleine Sharp, Alain Dagher, Edward A. Fon, D. Louis Collins, Quebec Parkinson Network

## Abstract

**Background:** Neuromelanin-MRI enables *in vivo* assessment of the substantia nigra (SN) and locus coeruleus (LC) in individuals with Parkinson’s disease (PD), yet longitudinal studies rely on cross-sectional processing that may introduce measurement variability and confound estimates of change over time.

**Objectives:** In this paper, a longitudinal neuromelanin-MRI processing framework is presented that is designed and validated to improve measurement stability and reduce processing-related variability across repeated scans.

**Methods:** Imaging and clinical data from the Quebec Parkinson Network were analyzed in 268 participants (199 PD, 69 controls), including a longitudinal subset of 74 participants (49 PD, 25 controls) scanned approximately one year apart. Validation experiments evaluated slice-by-slice intensity normalization for slice dependent intensity variation, bias field correction for LC signal asymmetry, and the effects of longitudinal registration on measurement stability and PD-control discrimination.

**Results:** Slice-by-slice intensity normalization significantly reduced brainstem intensity variability by 3.6%. A systematic leftward signal asymmetry was observed in the LC and persisted following N4 bias field correction, suggesting a scanner-related effect not captured by conventional bias field modeling. Longitudinal registration reduced annualized change variability by 25-36% for SN_CR_ and 27-34% for LC_CR_ metrics in controls, indicating improved within-subject measurement stability. Residual variability was also reduced for contrast-based metrics by up to 28%. Longitudinal registration generally produced larger PD-control effect sizes at baseline and follow-up, particularly for SN volume metrics. However, no significant *method × group × time* interactions were observed, indicating that estimated longitudinal trajectories did not differ significantly between longitudinal and conventional cross-sectional processing.

**Conclusions:** Longitudinal registration reduced technical variability and improved the precision of NM-MRI measurements. Although it did not significantly enhance detection of longitudinal PD-control differences over the follow-up interval examined here, it provides a more robust framework for longitudinal NM-MRI studies and may improve sensitivity to subtler biological effects in future investigations.

## 1.0 Introduction

The substantia nigra (SN) and locus coeruleus (LC) are key brainstem regions implicated in Parkinson’s disease. Both are bilateral, neuromelanin (NM)-rich nuclei, containing dopaminergic and noradrenergic neurons, respectively. The pigmented NM molecule accumulates in the SN and LC throughout much of the lifespan and may decline later in life (Chan-Palay & Asan, 1989; Zecca et al., 2001). In individuals with PD, NM is markedly reduced in the SN and LC due to neuronal degeneration. The SN and LC can be visualized *in vivo* using NM-MRI sequences such as 2D T1-weighted (T1w) turbo spin-echo (TSE) (Sasaki et al., 2006) and 3D gradient-echo (GRE) with magnetization transfer (Nakane et al., 2008), which exploit the paramagnetic properties and increased macromolecular content of NM relative to surrounding tissue. Cross-sectional NM-MRI studies consistently demonstrate reduced contrast- and volume-based NM-MRI measurements in the SN and LC in individuals with PD compared with healthy controls, establishing NM-MRI as a robust imaging biomarker for PD (He et al., 2023; Sulzer et al., 2018; Trujillo et al., 2024).

More recently, a limited number of longitudinal studies have used NM-MRI to quantify disease progression in PD. Matsuura et al. (2016) provided early evidence of longitudinal NM decline using T1w TSE NM-MRI in a small sample of 14 individuals with PD, quantifying NM using SN area and contrast ratio (CR) derived from semi-manual SN segmentation based on intensity thresholds. At follow-up, both total SN area and CR were reduced relative to baseline, with SN area declining in all participants.

Several later studies extended these observations and compared PD patients to control groups. Biondetti et al. (2020) measured NM-MRI volume and signal-to-noise ratios (SNRs) from manual SN segmentations in early and progressing PD and control cohorts. The study reported progressive, regionally specific SN NM loss in PD relative to controls, with earliest involvement observed in the sensorimotor SN. A follow-up study (Biondetti et al., 2021) described NM-MRI SNR changes across sensorimotor, associative, and limbic SN regions using automatic segmentation in template space. This work identified a spatiotemporal gradient of pathology with affected-side-related asymmetry; however, inter-visit changes in SN NM did not differ significantly between PD and controls. Gaurav et al. (2021) quantified NM-MRI in early and progressing PD cohorts against controls using both volume-based and signal intensity-based metrics derived from manual SN delineations, identifying baseline group differences. In this study’s longitudinal analysis, volume-based measurements declined significantly in PD but not in controls, with larger reductions observed in the progressing PD cohort, whereas contrast-based metrics were less reliable for detecting group differences over time.

An independent multisite study by Xing et al. (2022) examined longitudinal NM-MRI changes in PD and control groups using both manually derived contrast-based metrics (with explicit separation of the ventral and dorsal SN) and an automatic, template-based, normalized SN volume metric. Here, the PD group exhibited greater annualized NM loss for both contrast and volume metrics compared with controls, with the strongest effects observed in the ventral SN. Finally, Perot et al. (2025) examined longitudinal NM-MRI signal changes in PD and control groups, showing that after the age-related signal peak, NM-MRI signal decline is significantly accelerated in PD compared with the post-peak age-related decline observed in controls.

While existing longitudinal studies consistently support progressive NM loss, they differ in the reported sensitivity of contrast- and volume-based metrics and the magnitude of disease-related decline. One possible contributor to this variability is that all studies processed each imaging session independently (i.e., using cross-sectional processing), which may introduce measurement variability that obscures estimates of true biological change. In contrast, longitudinal processing jointly analyzes serial images from the same subject within a unified framework designed to improve the consistency and robustness of measurements across time.

Many of the studies mentioned rely on manual segmentation of the SN performed independently at each timepoint, with longitudinal change derived from separate measurements (Biondetti et al., 2020; Biondetti et al., 2021; Gaurav et al., 2021; Matsuura et al., 2016; Perot et al., 2025; Xing et al., 2022). Qualitative strategies such as side-by-side review of longitudinal scans or spatial smoothing have been used to reduce processing variability, but these do not replace an explicit longitudinal processing approach. Although some studies have reported high inter- and intra-rater reliability with manual segmentation, these assessments do not clearly evaluate within-subject longitudinal consistency. Some studies use template-based (Xing et al., 2022) and other automated (Hu et al., 2025) approaches which also process each visit independently, inferring longitudinal change from cross-sectional measurements. Collectively, these studies highlight the limited attention devoted to computational strategies that explicitly reduce within-subject measurement variability while measuring disease-related change.

Some noteworthy methodological efforts have been made to improve NM-MRI processing and quantification, including the work done by Wengler et al. (2020) that demonstrated high reproducibility of NM-MRI measurements and proposed automated processing procedures. However, their work focused on improving reproducibility and processing of GRE-based NM-MRI sequences, where explicit longitudinal registration and processing of T1w-TSE-based NM-MRI sequences were not mentioned. Hayat et al. (2022) developed a template-based framework aimed at facilitating harmonized quantification and multicenter implementation. However, this framework was primarily designed to improve spatial normalization across multicentre studies rather than to leverage information across timepoints through subject-specific longitudinal registration. Consequently, the impact of dedicated longitudinal processing strategies on NM-MRI measurement stability and sensitivity to disease-related change remains unclear.

The objective of this study was to develop and validate a longitudinal processing framework for 2D T1w TSE NM-MRI and determine whether dedicated longitudinal processing reduces measurement variability and improves the precision of quantitative NM-MRI measurements compared with conventional cross-sectional processing, while systematically evaluating additional processing strategies to minimize other sources of technical variability.

## 2.0 Methods

### 2.1 Data

#### Participants

Data used to develop and validate this pipeline framework were obtained from the Quebec Parkinson Network (QPN) (Gan-Or et al., 2020) database and included imaging as well as clinical assessments from 268 participants (199 PD, 69 age-matched controls). A subset of participants (49 PD, 25 controls) underwent follow-up imaging and clinical assessment after a median interval of 1.14 years (range: 0.94-3.97). Participants were recruited via neurologist referral or self-referral. Inclusion required age ≥18 years, proficiency in English or French, and capacity to provide informed consent. Exclusion criteria included other movement disorders, major neurological illness, prior neuroleptic use, and MRI contraindications. PD diagnoses were confirmed using a hierarchical certainty framework based on the MDS PD Diagnostic Criteria (Postuma et al., 2015), using neurologist confirmation, or patient self-report when neurologist verification was unavailable. PD participants receiving dopaminergic therapy were assessed in their usual ON state. All procedures were approved by the Research Ethics Board of the McGill University Health Centre, and all participants provided written informed consent under the Montreal Neurological Institute’s Open Science Clinical Biological Imaging And Genetic Repository (Das et al., 2022).

#### MRI Acquisition

All participants underwent a 60-minute imaging protocol in a 3T Siemens Prisma scanner equipped with a 32-channel head coil at the Montreal Neurological Institute. Details on the full protocol are provided in Bhagwat et al. (2025, under review). The sequences analyzed in this study included a 3D T1w MPRAGE sequence with a repetition time (TR) of 2300 ms, echo time (TE) of 2.98 ms, flip angle of 9°, field of view of 256 mm, and 1.0 mm isotropic voxel size. NM-MRI was acquired using a 2D T1w TSE sequence with TR = 600 ms, TE = 10 ms, flip angle = 120°, field of view of 220 mm, and a voxel size of 0.7 *×* 0.7 *×* 1.8 mm^3^. The 2D slice stack was aligned oblique-axially, with slice planes parallel to the anterior brainstem surface, centered on the superior pons, and extending inferiorly to include the fourth ventricle. The acquisition covered the SN and LC but not consistently the subcoeruleus complex, which was therefore excluded from the analysis. An example SN and LC from a representative subject is shown in Figure 1.

**Figure 1:**
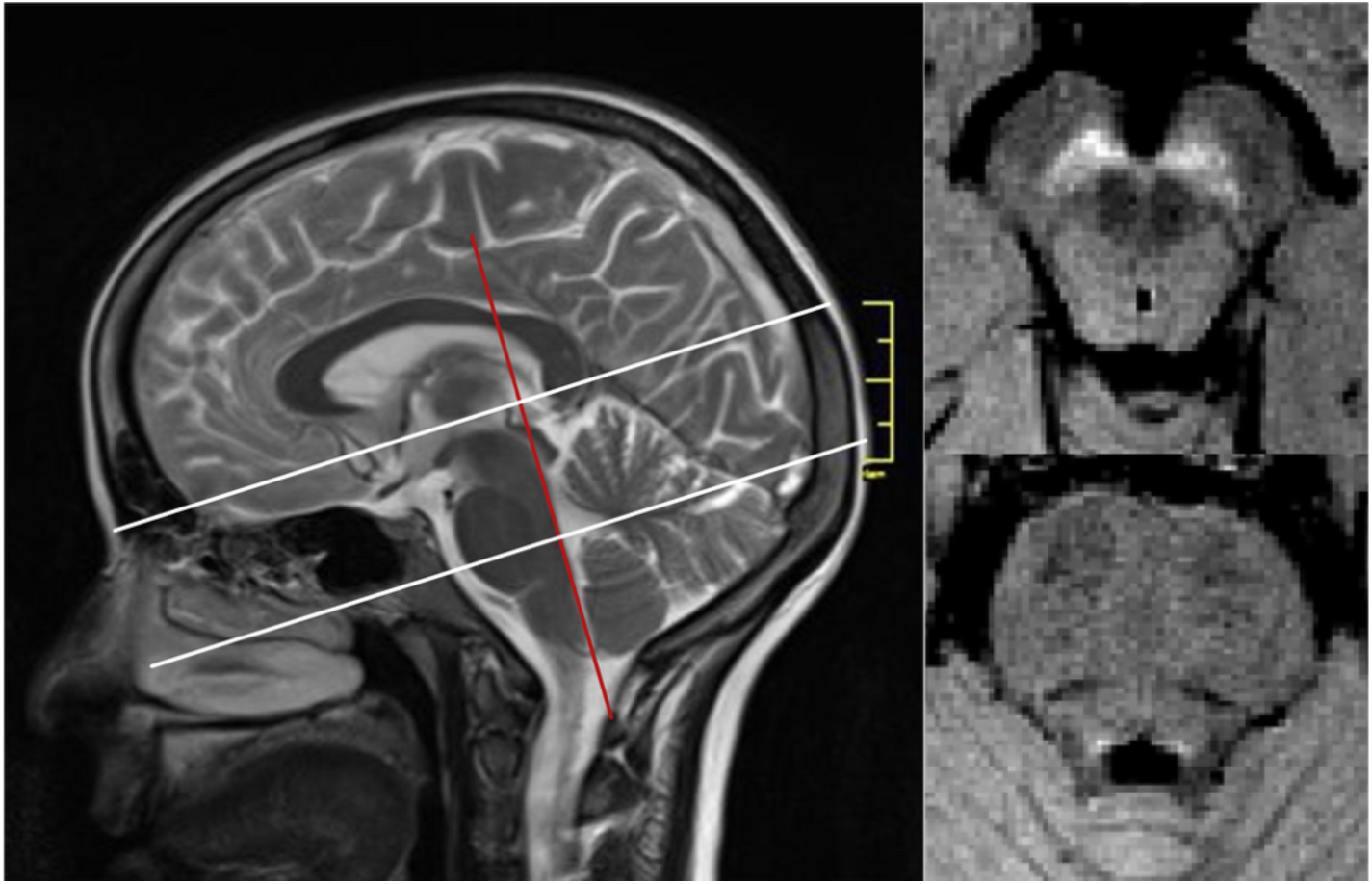
Imaging orientation and nuclei visualization. Sagittal view of 2D slice alignment and representative axial images of the SN and LC.

#### Clinical Assessments

Participants underwent standardized clinical assessments at each study visit. Both control participants and individuals with PD completed a neuropsychological assessment battery, including the Montreal Cognitive Assessment (MoCA). Individuals with PD additionally underwent the Movement Disorder Society Unified Parkinson’s Disease Rating Scale (MDS-UPDRS) and staging using the Hoehn and Yahr (H&Y) scale. Detailed descriptions of the cohort and assessment procedures are provided in Bhagwat et al. (2025, under review).

### 2.2 Neuromelanin Label Set Creation

A standardized label set was created to extract NM contrast and volume metrics from the SN and contrast metrics from the LC. All labels were defined using the symmetric PD126 stereotaxic template, a high-resolution multi-contrast MRI template derived from 126 individuals with PD (Madge et al., 2023b). All labels represented in PD126 space were symmetrized to avoid introducing geometric bias, as early-stage PD often presents unilaterally. The label set in PD126 stereotaxic space is shown as part of Figure 2.

**Figure 2.**
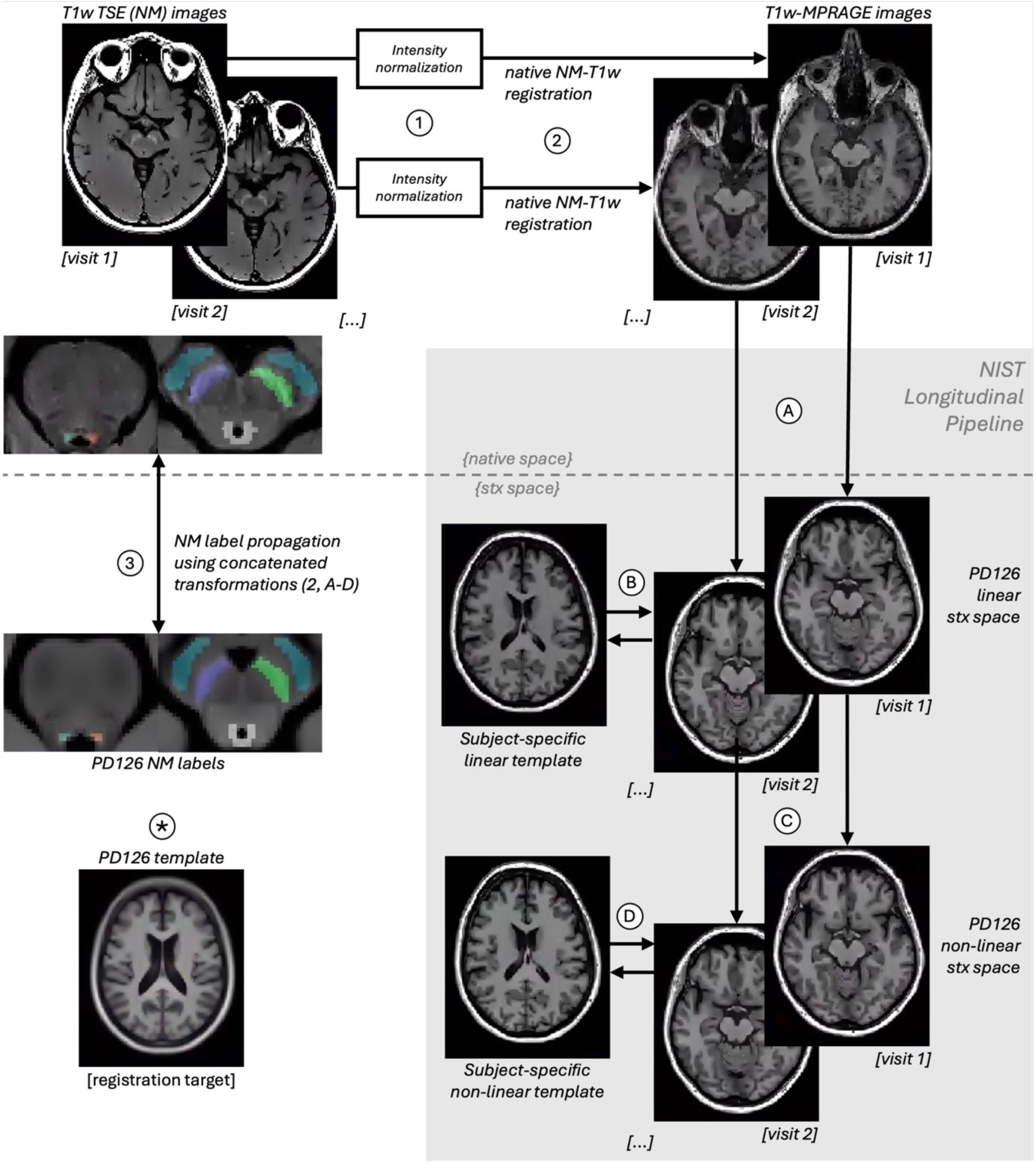
Longitudinal NM-MRI processing framework. Black arrows indicate registrations, and the dashed line separates native and stereotaxic (stx) processing. Native NM-MRI images undergo intensity normalization (1), before rigid registration to corresponding native T1w images (2). The NIST Longitudinal Pipeline (grey) calculates linear (A) and non-linear (C) stereotaxic transformations using the PD126 template (*) as the stereotaxic registration target. Subject-specific templates are created for subjects with multiple timepoints (visit 1, visit 2, …), generating additional transformations in linear (B) and non-linear (D) stx space that contribute to the final stereotaxic mapping. Atlas-defined NM labels in PD126 space are propagated to native NM-MRI space (3) using the inverse of the concatenated NM-to-T1w and longitudinal stereotaxic transformations (2, A-D). Labels for cerebral peduncles (blue), substantia nigra (indigo/lime green), locus coeruleus (red/green) and periaqueductal grey (white) are displayed in stx and native space. NM-MRI images may also be resampled to stereotaxic space using the same transformations in the forward direction.

SN functional subregion labels were obtained from Biondetti et al. (2021). As the source template was asymmetric, only the left-sided labels were non-linearly registered to PD126 using ANTs (Avants et al., 2008) and then reflected across the x-axis to generate right-sided labels. For the analyses in this study, all subregions within each SN were combined to form a single left ROI and a single right ROI. The full set of SN subregional labels was retained for use in other studies.

Probabilistic LC and periaqueductal gray (PAG) labels were obtained from the BrainstemNavigator package and aligned to PD126 following the recommended registration workflow (Bianciardi et al., 2015). Labels were symmetrized and thresholded to exclude cerebrospinal fluid while preserving potential LC or PAG contrast. LC volume was not quantified due to partial-volume effects at 3T resolution. A symmetric LC-shaped control region (LC-adj), shifted anteriorly and laterally, was created for LC signal asymmetry analyses.

For NM signal normalization, cerebral peduncle (CP) labels were manually delineated on one side of a study-specific in-house NM-MRI template, aligned to PD126, and then reflected across the x-axis to generate labels on the other side. The pontine tegmentum (PT) label (also used for NM signal normalization) was manually delineated directly in PD126 space, by an expert in neuroanatomy and image registration, and symmetrized. A brainstem mask encompassing SN, LC, CP, PAG and PT regions was generated from the symmetric PD126 NM volume and used to assess intensity homogeneity.

Labels used for NM-MRI quantification in this study (SN, LC, PAG, CP, and PT) underwent expert review by three neuroradiologists, who agreed that the labels accurately represented the corresponding anatomical structures.

### 2.3 Proposed Longitudinal NM-MRI Processing Framework

Figure 2 summarizes the proposed longitudinal NM-MRI processing framework used to quantify contrast- and volume-based metrics from the label set described above. The framework combines native space NM-MRI preprocessing with longitudinal T1w-derived registration to enable consistent label propagation and metric extraction across multiple timepoints. Candidate preprocessing steps aimed at reducing intensity-related variability and bias, together with the longitudinal registration strategy, were evaluated in the validation experiments described in Section 2.4.

The framework was implemented using the MINC 2.0 Toolkit (Vincent et al., 2016) with in-house Bash and Python (v3.12.7) scripts. The symmetric PD126 template served as the stereotaxic registration target for the entire framework.

Slice-by-slice intensity normalization (*inormalize)* from the MINC 2.0 Toolkit (Vincent et al., 2016) and N4 bias field correction (N4ITK) (Tustison et al., 2010) were implemented as candidate preprocessing steps to reduce intensity-related variability and bias in native space NM-MRI images.

To account for potential movement between scans, the NM-MRI images were rigidly co-registered to their corresponding T1w images in native space, using a midbrain bounding box to drive alignment by brainstem anatomy. This transform was concatenated with the T1w-derived longitudinal transformations to enable single-step label propagation from stereotaxic PD126 space to the native MRI space.

T1w-derived longitudinal transformations were acquired from T1w image processing using the NIST Longitudinal Pipeline (Aubert-Broche et al., 2013; Guizard et al., 2015). Briefly, T1w images underwent preprocessing in native space: brain extraction, denoising (Coupe et al., 2008), and intensity non-uniformity correction (Sled et al., 1998); followed by linear (Collins & Evans, 1997) and non-linear (Avants et al., 2008) registration to PD126 space. For longitudinal data, subject-specific linear and non-linear templates were generated to reduce variability in intra-subject measurements (Aubert-Broche et al., 2013; Guizard et al., 2015), and the resulting transformations were incorporated into the final stereotaxic mapping. All transformations were concatenated into a single native-to-stereotaxic warp per timepoint to minimize resampling-related blurring. The most recent version of the pipeline can be found here: https://github.com/NIST-MNI/nist_mni_pipelines.

Once resampled to native NM-MRI space, the NM label set was used to extract contrast- and volume-based metrics from the NM-MRI images. Contrast-based metrics were computed as contrast ratios for the SN and LC using Equation 1:

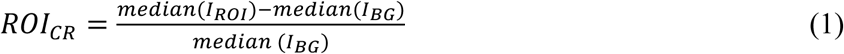

Where *ROI_CR_* is the contrast ratio for SN or LC, *I_R_*_O*I*_ represents the intensity distribution within the target ROI, and *I_BG_* represents the intensity distribution within the corresponding background ROI. Median intensity values were used to mitigate any potential skewed distributions. The CP served as the background ROI for SN_CR_, while the PT and PAG were used as candidate background ROIs for LC_CR_, and are later evaluated in a small validation experiment described in Section 2.4.

Raw SN volume was calculated as the cumulative volume of suprathreshold voxels within the SN ROI:

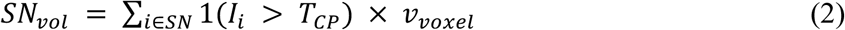

where *SN_vol_* is the raw SN volume, 1(·) is an indicator function that equals 1 when voxel intensity exceeds the threshold and 0 otherwise, *I_i_* is the intensity of each voxel inside the SN ROI, *T_CP_* is the threshold derived from CP intensity, and *v_voxel_* is the voxel volume (mm^3^). The threshold identifies SN voxels exhibiting NM signal greater than background CP intensity and was defined as:

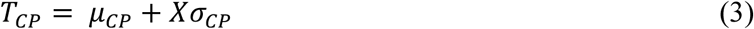

where *μ_CP_* and *σ_CP_* are the mean and standard deviation of CP intensity, respectively, and *X* is the multiplier. Following Xing et al. (2022), the multiplier was selected to produce SN volume estimates comparable to postmortem measurements of normally pigmented SN tissue (Pakkenberg et al., 1991). Normalized SN_vol_ is obtained by dividing the raw SN_vol_ by the subject’s intracranial capacity (ICC), estimated as the sum of grey matter, white matter, and cerebrospinal fluid volumes measured from the NIST Longitudinal Pipeline (Aubert-Broche et al., 2013; Guizard et al., 2015). Because SN volume is derived from intensity thresholding within the SN ROI, it reflects the volume of NM-positive tissue rather than the anatomical volume of the SN itself. Raw SN_vol_ is included for comparison to previous longitudinal studies (Biondetti et al., 2020; Gaurav et al., 2021).

All NM-MRI contrast- and volume-based metrics were computed separately for left and right nuclei. Labels could alternatively also be retained in stereotaxic space, when NM-MRI images are required in standard space for group analyses.

### 2.4 Experiments

#### Evaluation of Slice-by-Slice Intensity Normalization

T1w TSE NM-MRI can exhibit slice dependent intensity variation due to non-uniform excitation and residual RF cross-talk between slices, despite interleaving strategies intended to reduce these striping artifacts (Zun et al., 2014). A validation experiment was conducted to evaluate whether slice-by-slice intensity normalization reduces brainstem intensity variability.

Control group NM-MRI images were used for this experiment and were treated as independent scans. For each scan, two preprocessing states were defined: “before normalization”, corresponding to the raw image, and “after normalization”, corresponding to the image following slice-by-slice intensity normalization. For the latter state, normalization was applied along the z-direction to the entire image volume using the *inormalize* preprocessing step. By default, *inormalize* estimates the median ratio of voxel intensities between adjacent slices and rescales each slice accordingly to reduce slice-to-slice intensity variation. Using *inormalize* ensured consistency with the MINC-based NM-MRI processing pipeline and avoided unnecessary resampling or format conversion.

For each scan and preprocessing state, voxel intensities were extracted from a brainstem mask defined on the PD126 template and warped to native space. Because the SN, LC and reference ROIs are located within the brainstem, intensity variability within the brainstem was quantified as the standard deviation (SD) of all brainstem voxels.

#### Evaluation of Bias Field Correction for LC Signal Asymmetry

Scanner-dependent left-right signal asymmetry has been reported in the LC in NM-MRI studies, with direction varying across scanner platforms, and has been attributed to spatial variations in the B₁ field rather than biological lateralization (Betts et al., 2017; Trujillo et al., 2024). A validation experiment was performed to quantify left-right signal asymmetry in NM-MRI data and to evaluate whether a bias field correction preprocessing step mitigates this effect. The objectives were to (i) assess signal asymmetry in the LC, (ii) determine whether a similar scanner-dependent signal asymmetry is present in the SN, (iii) evaluate signal asymmetry in adjacent non-NM tissue, and (iv) test the effect of a bias field correction preprocessing step across these ROIs.

Control group NM-MRI images were used for this experiment and treated as independent scans, assuming all ROI signal asymmetry reflects a systematic scanner-dependent effect. For each scan, two preprocessing states were defined: “before correction”, corresponding to the raw image, and “after correction”, corresponding to the image following bias field correction. N4ITK bias field correction was applied in 3D with intensity rescaling, a B-spline grid spacing of 120 mm, a shrink factor of 2, and 200 × 200 × 200 iterations with a convergence threshold of 1 × 10⁻⁷. Parameters were selected to model the hypothesized broad B_1_ field variation while preserving spatial detail in the bilateral LC.

For each scan and preprocessing state, median left and right ROI intensities were extracted from the LC and SN, as well as from two non-NM regions: an LC shape-matched region adjacent to the LC (LC-adj), and the CP. Left-right signal asymmetry was quantified using a symmetry index (SI), calculated as (I_left_ - I_right_) / (I_left_ + I_right_), where *I_x_* is the ROI median intensity for side *x*. Reductions in |SI| “after correction” were interpreted as decreased scanner-related signal asymmetry. To support this interpretation, N4-estimated bias fields were visually inspected for left-right gradients in the LC. Because B₁ field maps were not acquired in the QPN imaging protocol, B₁ field maps from an independent study on the same scanner were examined to assess left-right variation in the vicinity of the LC as a potential source of the observed signal asymmetry. Finally, LC signal asymmetry was assessed in the PD cohort to evaluate whether bias characterization was consistent across groups.

#### Validation of Longitudinal Registration for Within-Subject Stability

Longitudinal NM-MRI studies that use cross-sectional processing are sensitive to registration and segmentation inconsistencies across timepoints, which can introduce artificial within-subject variability and bias estimates of biological change in the SN and LC. A validation experiment was conducted to evaluate whether longitudinal registration improves the stability of NM-MRI contrast and volume-based metrics relative to conventional cross-sectional processing.

Longitudinal NM-MRI data from the control group were processed using two registration approaches: a “cross-sectional” approach and a “longitudinal” approach. Both approaches included the rigid co-registration warp of the NM-MRI scan to its corresponding T1w scan from the same timepoint, as well as linear and nonlinear transformations to the PD126 template, while the “longitudinal” approach additionally incorporated transformations for the linear and non-linear subject-specific longitudinal templates. Based on the results from the first validation experiment, *inormalize* was applied as the intensity normalization preprocessing step before both approaches, and pooled CP labels were used when extracting background median intensities and when deriving volume thresholds. No additional compensatory preprocessing based on the findings of the second validation experiment was applied here, as doing so could have confounded the comparison between the two registration approaches.

Registration performance was evaluated primarily using within-subject annualized change of NM-MRI metrics in the control group, where minimal biological change was expected across visits. Annualized change was calculated as the difference between follow-up and baseline metrics divided by the time, in years, between scans. Comparing the variance of annualized change for each metric measured for the control group, derived from cross-sectional versus longitudinal approaches, facilitated assessment of whether longitudinal processing reduced the variability of apparent within-subject fluctuations.

#### Validation of Longitudinal Registration for Improved Effect Size

A validation experiment was performed to assess whether the anticipated within-subject stability from longitudinal registration translates to significantly larger effect sizes for group differences in NM-MRI metrics, relative to conventional cross-sectional processing.

Both PD and control groups were used for this experiment. Longitudinal NM-MRI data from the PD group were processed identically to the control group, including all intensity normalization and metric quantification steps, with NM-MRI metrics derived using both “cross-sectional” and “longitudinal” registration approaches. To improve model statistical power and strengthen baseline estimates, NM-MRI data from subjects with only a baseline visit were also included in the analysis.

To determine whether longitudinal registration improved the ability of NM-MRI metrics to distinguish PD from controls, mixed-effects models were used to compare PD-control group separation between cross-sectional and longitudinal registration approaches by testing *group × method* and *group × method × time* interactions for each metric. These analyses assessed whether longitudinal registration produced larger PD-control effect sizes and improved detection of longitudinal NM-MRI change relative to the conventional cross-sectional registration approach. In addition to evaluating group separation, residual variability was compared between registration approaches to assess whether longitudinal registration improved measurement consistency across NM-MRI metrics. Details on the models used and how the residual variance of each method within the model was calculated is explained further in the following section.

### 2.5 Statistical analysis

All statistical analyses were performed in R (version 4.3.2). Normality was assessed using the Shapiro-Wilk test. A significance threshold of α = 0.05 was applied to all analyses.

Group differences in demographic and clinical variables were evaluated using independent samples t-tests or Wilcoxon rank-sum tests, as appropriate; sex distributions were compared using chi-square tests.

#### Evaluation of slice-by-slice intensity normalization

A Wilcoxon signed-rank test was used to compare SD measurements of brainstem voxel intensities before and after slice-by-slice normalization to assess whether the preprocessing step reduced brainstem intensity variability.

#### Evaluation of bias field correction for LC signal asymmetry

Left-right signal differences were evaluated using paired t-tests or Wilcoxon signed-rank tests in each ROI before and after bias field correction (LC, LC-adj, SN, CP). Reductions in scanner-related asymmetry were tested using paired t-tests or Wilcoxon signed-rank tests on |SI| values. Group differences in LC signal asymmetry between control and PD groups were analyzed using Welch two-sample t-tests or Wilcoxon rank-sum tests, as appropriate.

#### Validation of longitudinal registration for within-subject stability

For each NM-MRI metric, the variance of annualized change was compared between metric values derived using the cross-sectional and longitudinal registration approaches. Comparisons were performed using Pitman-Morgan tests, or paired permutation tests when normality assumptions were not met.

#### Validation of longitudinal registration for improved effect size

Linear mixed-effects models were used to evaluate whether longitudinal registration improved PD-control group separation relative to the conventional cross-sectional registration approach. Models were fit separately for each NM-MRI metric using longitudinal and baseline-only data, with fixed effects for the time between visits, the method used (cross-sectional vs longitudinal registration), the group (PD vs. controls) and all interaction terms, with a subject random intercept to account for repeated measurements. The primary model equation, shown in Equation 4, was:

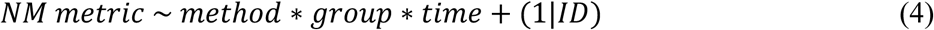

where *method* represented the registration approach (cross-sectional vs longitudinal), *group* represented diagnostic group (PD vs control), *time* represented the time since baseline, and +(1|*ID*) was the subject random intercept. Baseline-only participants were incorporated by assigning *time* = 0, allowing inclusion of subjects with a single visit to improve estimation of baseline group differences and overall model stability.

The *method × group* interaction term was used to assess whether PD-control group separation differed between registration approaches at baseline, while *method × group × time* interaction term was used to assess whether the longitudinal registration approach improved detection of PD-control differences over time relative to the conventional cross-sectional registration approach.

Residual variance was compared between registration approaches, as lower unexplained variability may indicate reduced processing-related noise. For each NM-MRI metric, mixed-effects models allowing separate residual variances for cross-sectional and longitudinal registration methods were compared with primary models, which assumed a common residual variance across methods. These comparisons assessed whether residual variability differed significantly between registration approaches. Method-specific residual variance estimates were subsequently compared descriptively to determine whether longitudinal registration was associated with lower unexplained variability than cross-sectional registration. Separate residual variance structures were evaluated as a secondary analysis rather than incorporated into the primary models to preserve a common variance structure for consistent PD-control comparisons between registration approaches.

P-values for fixed effects in the primary linear mixed-effects models were obtained using Wald t-tests with Satterthwaite approximation of degrees of freedom. Likelihood ratio tests were used for comparisons between pooled-variance and method-specific residual variance models. False discovery rate (Benjamini-Hochberg) correction was applied across metrics within each analysis.

## 3.0 Results

Baseline and follow-up demographic and clinical assessment information from the QPN PD and control groups are summarized in Table 1. At both baseline and follow-up, the PD and control groups did not differ significantly in age at MRI acquisition or years of education. Sex distribution differed significantly between groups at both time points, with a higher proportion of males in the PD cohort. Controls scored significantly higher than individuals with PD on the MoCA at both baseline and follow-up assessments. Clinical measures specific to PD, including disease duration, MDS-UPDRS III, and Hoehn & Yahr stage, are reported for the patient group only at each time point.

**Table 1:** Baseline and follow-up demographic and clinical assessment information for all QPN participants. Values shown as mean ± SD (range min-max). P-values indicate group comparisons.

|  | Baseline |  |  | Follow-up |  |  |
| --- | --- | --- | --- | --- | --- | --- |
|  | PD<br>(n=199) | Controls<br>(n= 69) | p-value | PD<br>(n=49) | Controls<br>(n= 25) | p-value |
| **Demographics** |  |  |  |  |  |  |
| Age at MRI acquisition (years) | 65.3 ± 8.9<br>(43.3 - 88.3) | 62.6 ± 11.9<br>(26 - 83) | p = 0.2461<sup>†</sup> | 68.0 ± 7.9<br>(51.4 - 89.3) | 66.2 ± 8.8<br>(44.9 - 81.8) | p = 0.3931 |
| Sex, male/female | 133 / 66 | 25 / 44 | p < 0.001<sup>‡</sup> | 35 / 14 | 7 / 18 | p < 0.001<sup>‡</sup> |
| Years of education | 12.4 ± 7.3<br>(<6 - 20+) | 14.9 ± 1.5<br>(6 - 19) | p = 0.5254<sup>†</sup> | 14.2 ± 3.6<br>(6 - 20) | 15.5 ± 2.7<br>(11 - 19) | p = 0.175 |
| **Clinical Scores** |  |  |  |  |  |  |
| MoCA | 25.2 ± 3.3<br>(14 - 30) | 26.7 ± 2.7<br>(16 - 30) | p < 0.005<sup>†</sup> | 25.6 ± 2.86<br>(18 - 30) | 27.6 ± 1.41<br>(25- 30) | p < 0.005<sup>†</sup> |
| Disease duration | 5.0 ± 4.5<br>(0.07 - 25.1) | N/A | N/A | 5.4 ± 3.6<br>(1.3 - 19.3) | N/A | N/A |
| MDS-UPDRS III | 29.6 ± 13.6<br>(5 - 72) | N/A | N/A | 34.8 ± 11.7<br>(10 - 65) | N/A | N/A |
| Hoehn & Yahr stage | 1.8 ± 0.6<br>(1 - 4) | N/A | N/A | 2.15 ± 0.4<br>(1 - 2) | N/A | N/A |

<sup>†</sup> Wilcoxon rank-sum. <sup>‡</sup>Chi-square. N/A = Not applicable.

### Evaluation of Slice-by-Slice Intensity Normalization

A total of 89 control scans were included in the analysis (64 baseline and 25 follow-up). Five baseline scans were excluded following quality control failure from flow artefact, movement artefact, or misregistration. No visible cross-talk artefacts were identified during quality control of the study images prior to slice-by-slice normalization; however, residual slice-dependent intensity variation consistent with non-uniform excitation was observed. Figure 3 illustrates the effect of slice-by-slice normalization on an external dataset exhibiting visible cross-talk artefacts. In the QPN control cohort, slice-by-slice normalization significantly reduced brainstem intensity variability across scans by 3.6% (Wilcoxon signed-rank test: V = 83, p < 0.0001; Figure 4).

**Figure 3.**
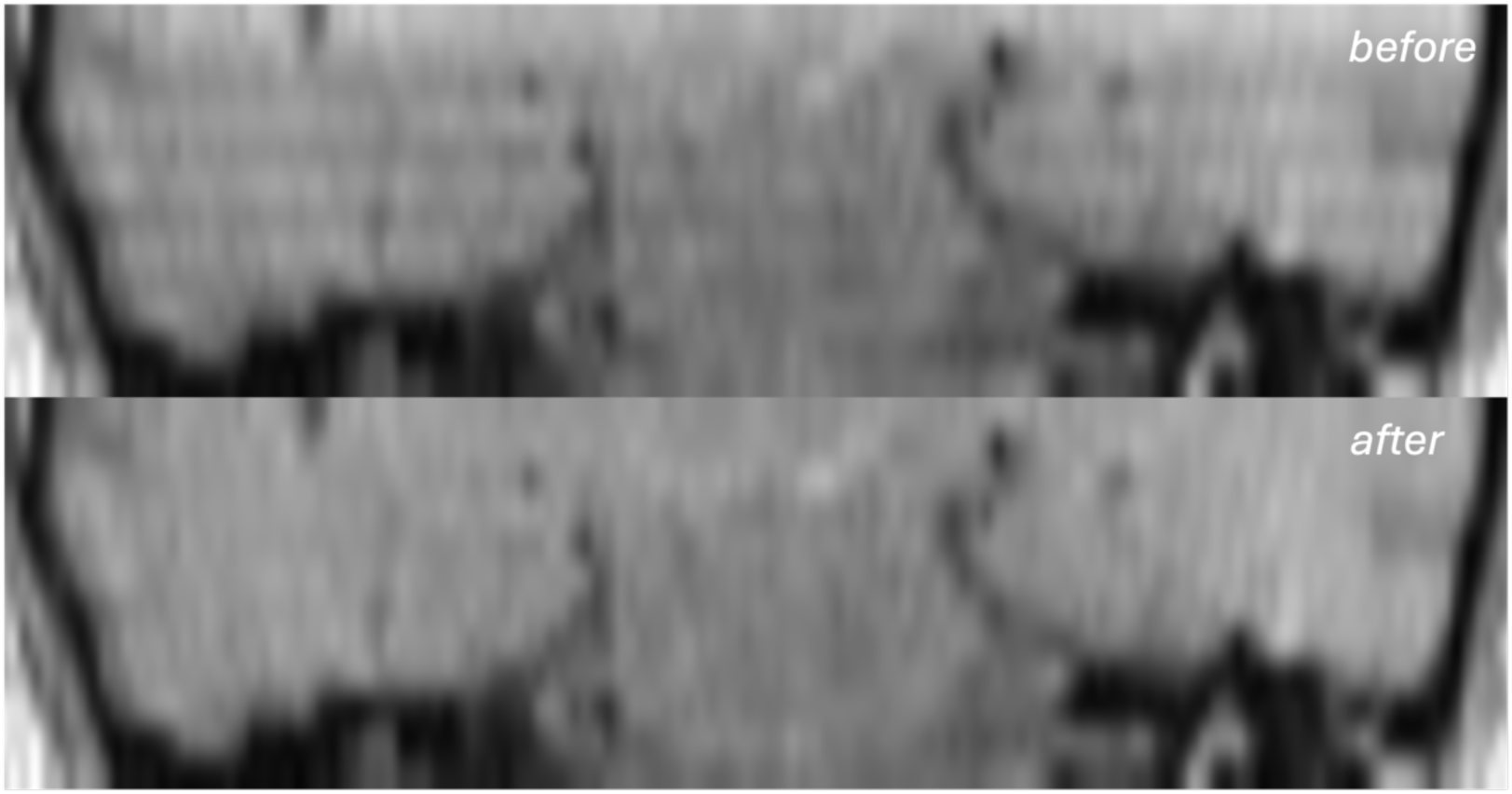
Effect of slice-by-slice normalization on external dataset with cross-talk artefact. Coronal slice shown before (top) and after (bottom) slice-by-slice normalization of a transverse acquisition. Slice-wise striping consistent with cross-talk is present before normalization and absent after normalization. The external dataset is shown solely to illustrate the effect of the algorithm under conditions where striping artefacts are visually apparent.

**Figure 4.**
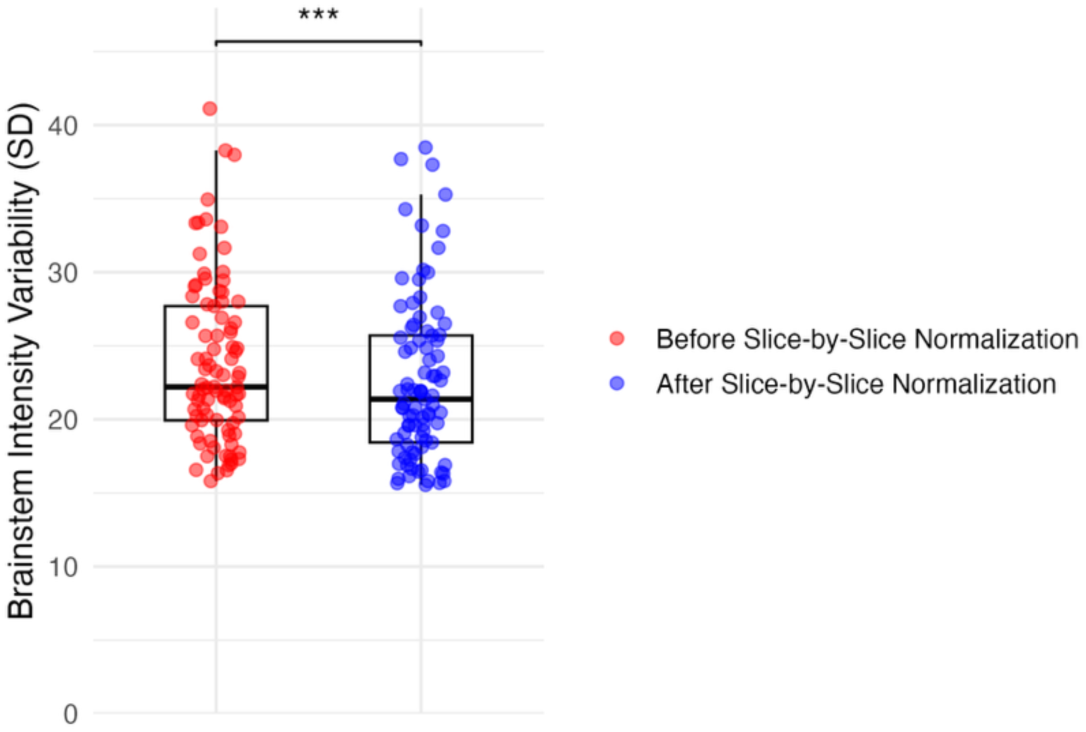
Effect of slice-by-slice normalization on brainstem intensity variability. Voxelwise brainstem intensity SDs are shown before (red) and after (blue) slice-by-slice normalization. Points represent individual scans from the control group. *** indicates a significant difference (p < 0.001) between pre- and post-normalization brainstem intensity SDs.

### Evaluation of Bias Field Correction for LC Signal Asymmetry

A total of 89 control scans were included in the analysis (64 baseline and 25 follow-up), consistent with the previous experiment. In the control cohort, significant leftward asymmetry (i.e., greater NM signal on the left) was observed in both the LC (1.8%) and the LC-adjacent region (1.5%) before bias field correction and persisted following correction (see Table 2 and Fig. 5). The CP exhibited significant rightward asymmetry before and after correction (0.16% and 0.15%), whereas the SN did not show significant asymmetry prior to correction but demonstrated a small but significant leftward bias (0.24%) after correction. A significant reduction in |SI| was found only for the CP with bias field correction, while changes in the LC, adjacent LC, and SN regions did not reach significance.

**Figure 5.**
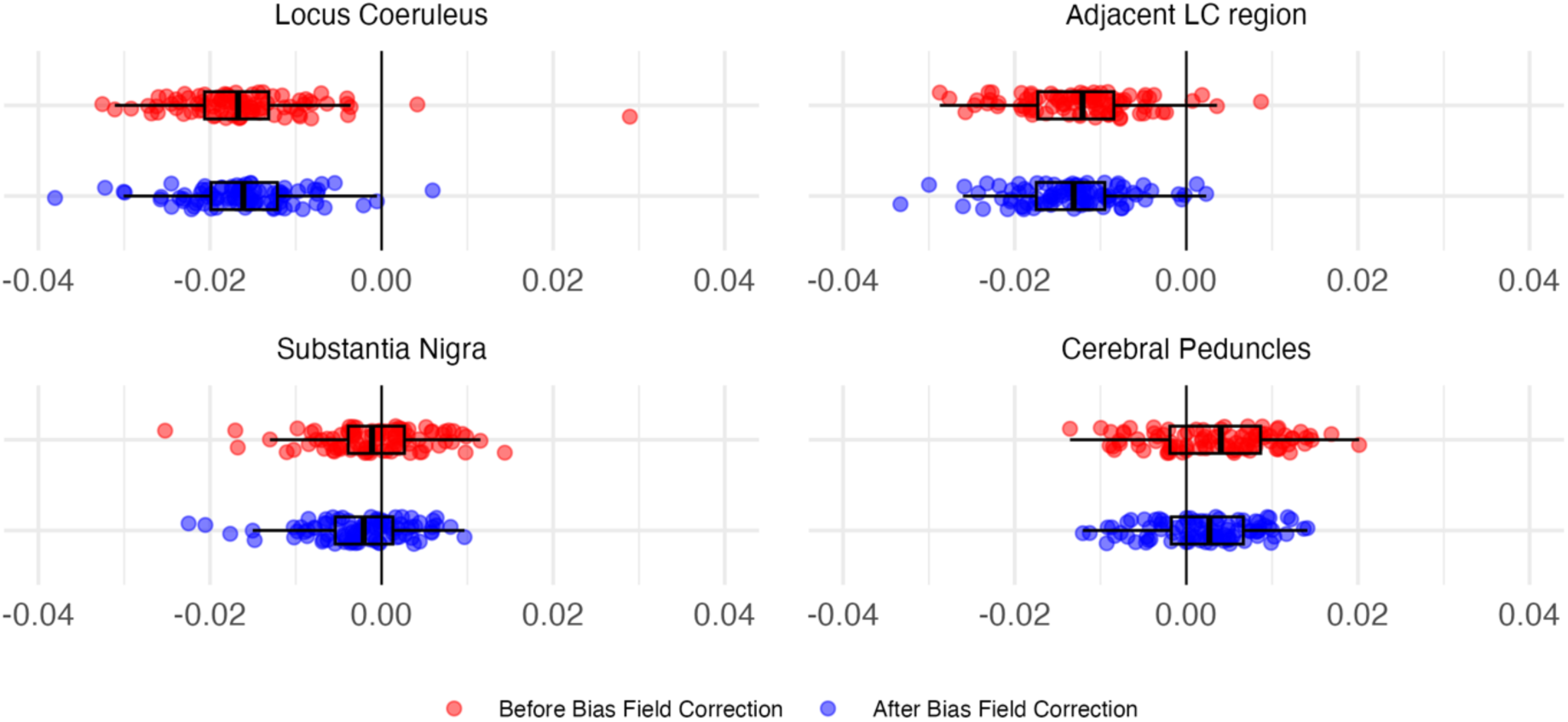
Symmetry indices of the LC (top left), a non-NM region adjacent to the LC (top right), SN (bottom left), and CP (bottom right) in control-group images before (red) and after (blue) applying bias field correction. Negative values indicate a higher intensity on the left. Note that median NM-MRI intensity values ranged from 189-433 in the LC, 151-411 in LC-adj, 210-446 in the SN, and 204-386 in the CP

**Table 2.**
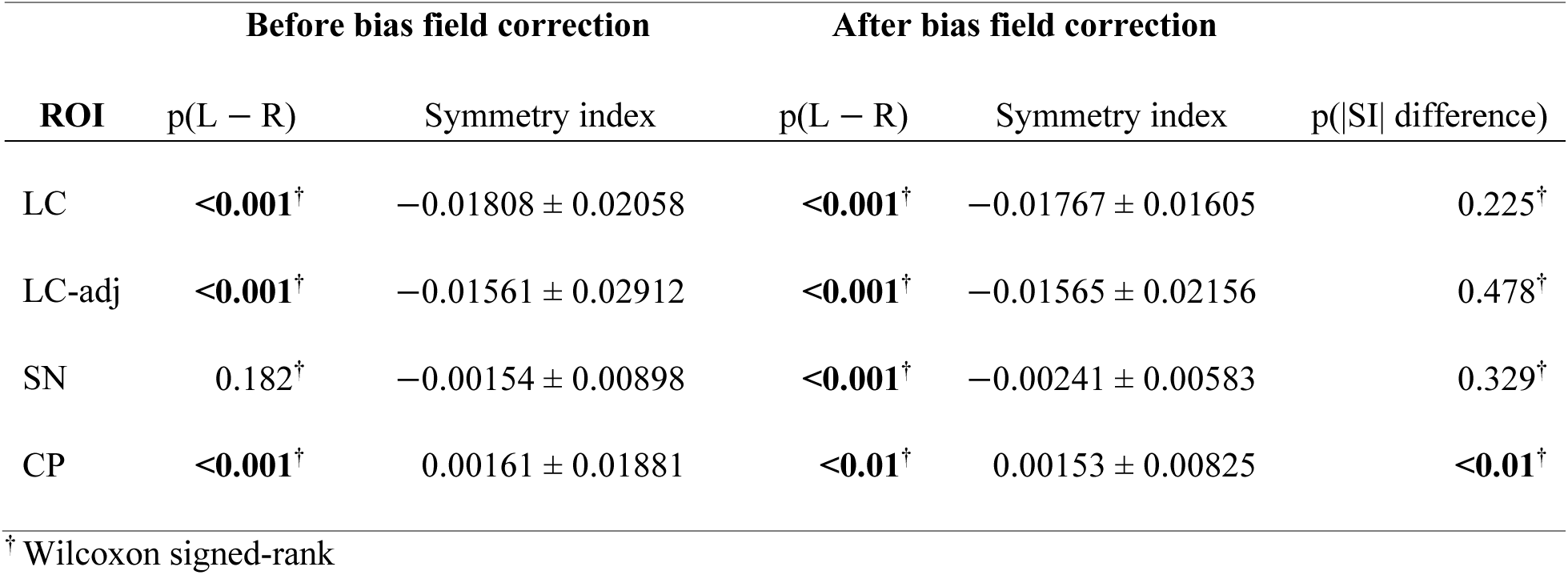
Left-right asymmetry before and after bias field correction across LC, LC-adj, SN, and CP regions. Reported values include the significance of left-right differences (p-values), mean symmetry index (mean ± SD), and the p-value difference in |SI| following correction. Bolded p-values indicate significance.

Further inspection of the N4-estimated bias fields in the current dataset showed smooth and low-frequency correction fields across all subjects, with no apparent left-right gradient at the level of the bilateral LC.

In an independent dataset acquired on the same scanner (n = 10, MPRAGE acquisition), B₁ field maps demonstrated a spatial bias in the LC region (1.7% ± 3.6%), with the orientation of this bias directed rightward.

There was no significant difference in LC SI between the PD and control groups (Wilcoxon rank-sum test: W = 10 708, p = 0.742; Figure 6).

**Figure 6:**
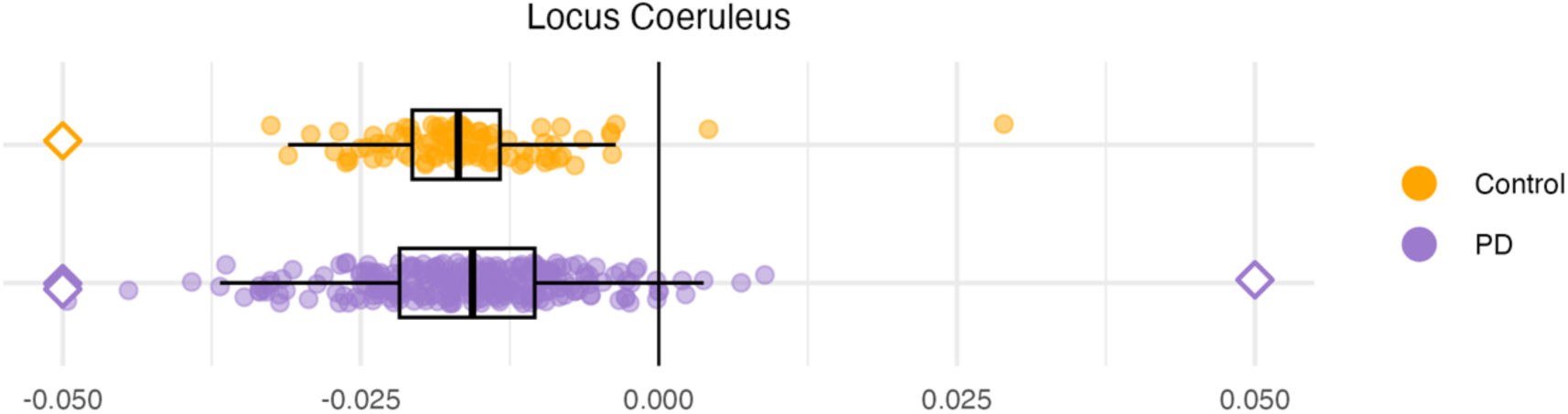
Symmetry indices of the LC in PD and control participants. Outliers beyond the display bounds are plotted as capped diamonds at the axis edges.

### Validation of Longitudinal Registration for Within-Subject Stability

A total of 24 control subjects were included in this analysis. Longitudinal data from one control subject was excluded due to image artefact from one timepoint affecting registration performance.

Assessing within-subject longitudinal stability in the control group, annualized change SDs were reduced when metrics were derived using the longitudinal (LNG) approach compared with the cross-sectional (XS) approach (Table 3). For SN_CR_, the longitudinal approach reduced annualized change SD by 25.1% (left) and 36.4% (right), with both reductions reaching statistical significance.

**Table 3.** Longitudinal stability comparison of NM-MRI metrics in controls processed using cross-sectional (XS) and longitudinal (LNG) registration approaches. Stability was assessed by measuring the variability of annualized change between baseline and follow-up visits. LNG reduction (%) represents the percentage reduction in annualized-change SD achieved using the longitudinal registration approach relative to XS registration. P-values test whether the variance of annualized change differed significantly between registration approaches. Note that annualized change SDs are shown for interpretability, while statistical comparisons were performed on variances. Average baseline metric values are shown to facilitate interpretation of the annualized change SDs relative to the scale of each metric.

| metric | Cross-sectional reg. |  | Longitudinal reg. |  | LNG Red.<br>(%) | p-value |
| --- | --- | --- | --- | --- | --- | --- |
| | Mean<br>(t = 0) | $\Delta$ SD | Mean<br>(t = 0) | $\Delta$ SD | | |
| $\Delta SN_{CR}$ (Left) | 0.14602 | 0.00929 | 0.15014 | 0.00695 | **25.13** | **<0.05** |
| $\Delta SN_{CR}$ (Right) | 0.14300 | 0.01154 | 0.14734 | 0.00735 | **36.36** | **<0.001** |
| $\Delta$ raw $SN_{vol}$ (Left) | 117.37 | 20.67 | 131.45 | 19.74 | 4.52 | 0.528<sup>†</sup> |
| $\Delta$ raw $SN_{vol}$ (Right) | 114.29 | 21.57 | 130.24 | 19.11 | 11.40 | 0.087<sup>†</sup> |
| $\Delta$ norm $SN_{vol}$ (Left) | 8.27e-05 | 1.47e-05 | 9.28e-05 | 1.41e-05 | 4.13 | 0.553<sup>†</sup> |
| $\Delta$ norm $SN_{vol}$ (Right) | 8.04e-05 | 1.51e-05 | 9.17e-05 | 1.35e-05 | 10.81 | 0.077<sup>†</sup> |
| $\Delta LC_{CR\_PAG}$ (Left) | 0.01387 | 0.01577 | 0.02371 | 0.01151 | **27.02** | **<0.05** |
| $\Delta LC_{CR\_PAG}$ (Right) | -0.01866 | 0.01492 | -0.01228 | 0.00983 | **34.15** | **<0.01** |
| $\Delta$ LC<sub>CR_PT</sub> (Left) | 0.03364 | 0.01476 | 0.04094 | 0.01383 | 6.29 | 0.702 |
| $\Delta$ LC<sub>CR_PT</sub> (Right) | 0.00046 | 0.01493 | 0.00434 | 0.01154 | 22.73 | 0.108 |

<sup>†</sup> paired permutation testing

For raw SN_vol_, the longitudinal approach reduced annualized change SD by 4.5% (left) and 11.4% (right), although neither reduction was significant. Similarly, for normalized SN_vol_, the longitudinal approach reduced annualized change SD by 4.1% (left) and 10.8% (right), but neither reduction was significant. For LC_CR_PAG_, the longitudinal approach reduced annualized change SD by 27.0% (left) and 34.2% (right), with both reductions reaching statistical significance. In contrast, LC_CR_PT_ showed smaller and non-significant reductions in annualized change SD. From these results, PAG demonstrated greater longitudinal stability than PT in this dataset and was therefore selected as the LC reference region for subsequent analyses.

### Validation of Longitudinal Registration for Improved Effect Size

A total of 46 PD subjects were included in this analysis along with the controls from the previous analysis. Longitudinal data from 2 PD subjects were excluded following quality control failure due to image artifacts from one of the timepoints affecting registration performance. Table 4 summarizes the results of the linear mixed-effects models. Significant *group* effects were observed for bilateral SN contrast and volume-based metrics under the reference cross-sectional registration approach. In contrast, no significant overall group effects were observed for LC_CR_ metrics. Significant *method* effects were observed for all NM-MRI metrics (Table 4), denoting systematically different NM-MRI measurements between cross-sectional and longitudinal registration approaches. Significant *method × group* interactions were observed only for bilateral SN_vol_ metrics (raw and normalized), denoting that PD-control differences varied between registration approaches at the reference time point (baseline).

**Table 4:**
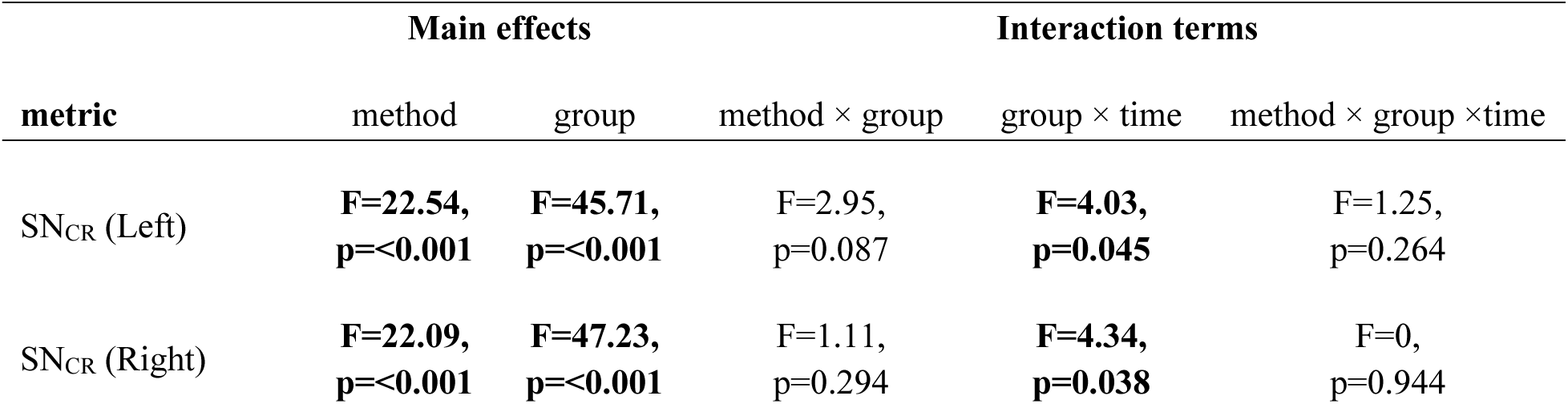
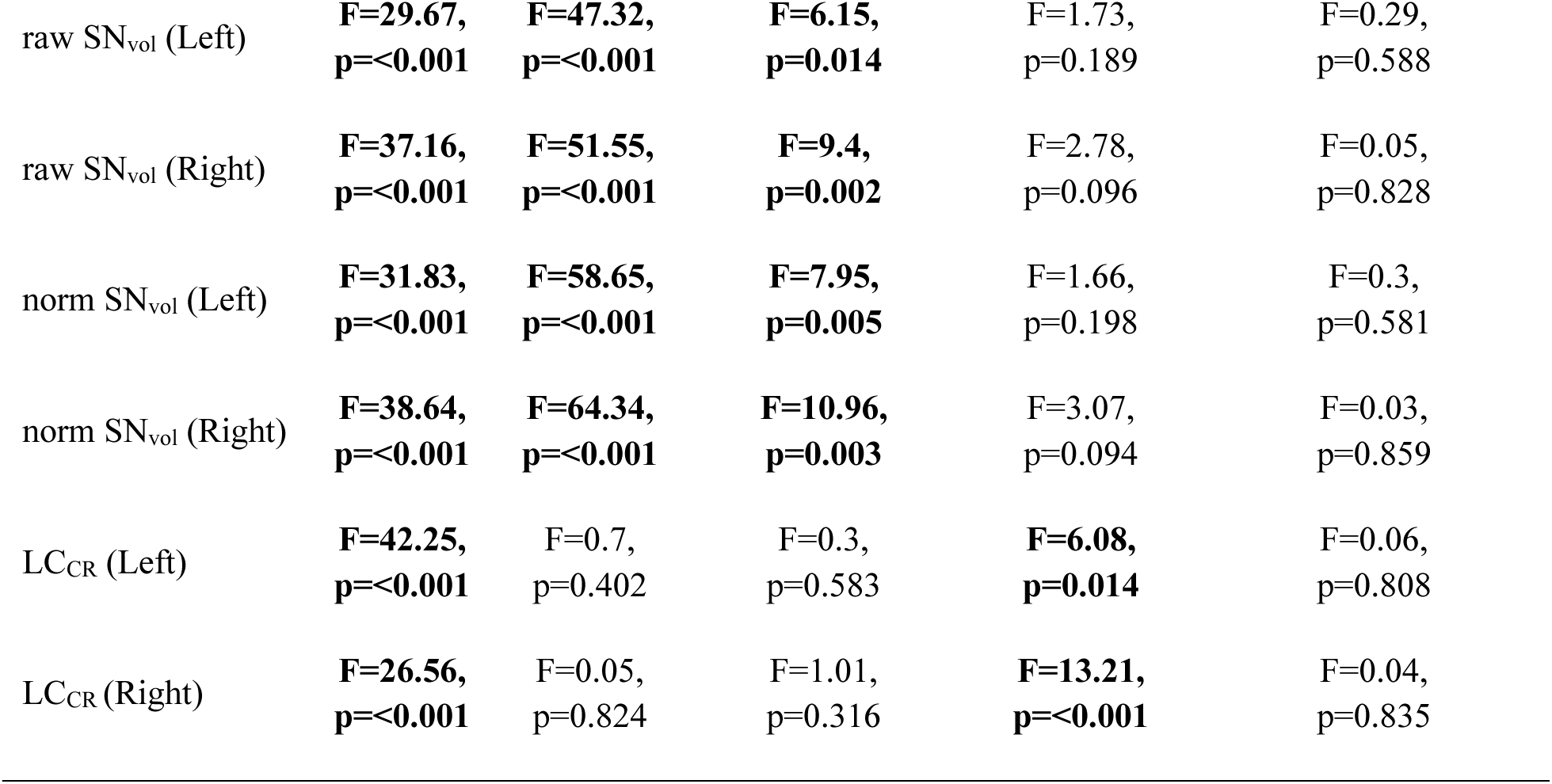
Results of linear mixed-effects models assessing the effects of registration method (XS vs. LNG), diagnostic group (PD vs. control), and time between baseline and follow-up scans on NM-MRI metrics. F-statistics and corresponding p-values are shown for the main effects of method and group, and for the method × group, group × time, and method × group × time interaction terms. Significant results (p < 0.05) are bolded.

Significant *group × time* interactions were observed for bilateral SN_CR_ and bilateral LC_CR_ metrics, indicating differing rates of change between groups under the reference cross-sectional registration approach. However, no significant *method × group × time* interactions were detected for any metric, indicating that the estimated longitudinal trajectories did not differ significantly between registration approaches (Figure 7).

**Figure 7.**
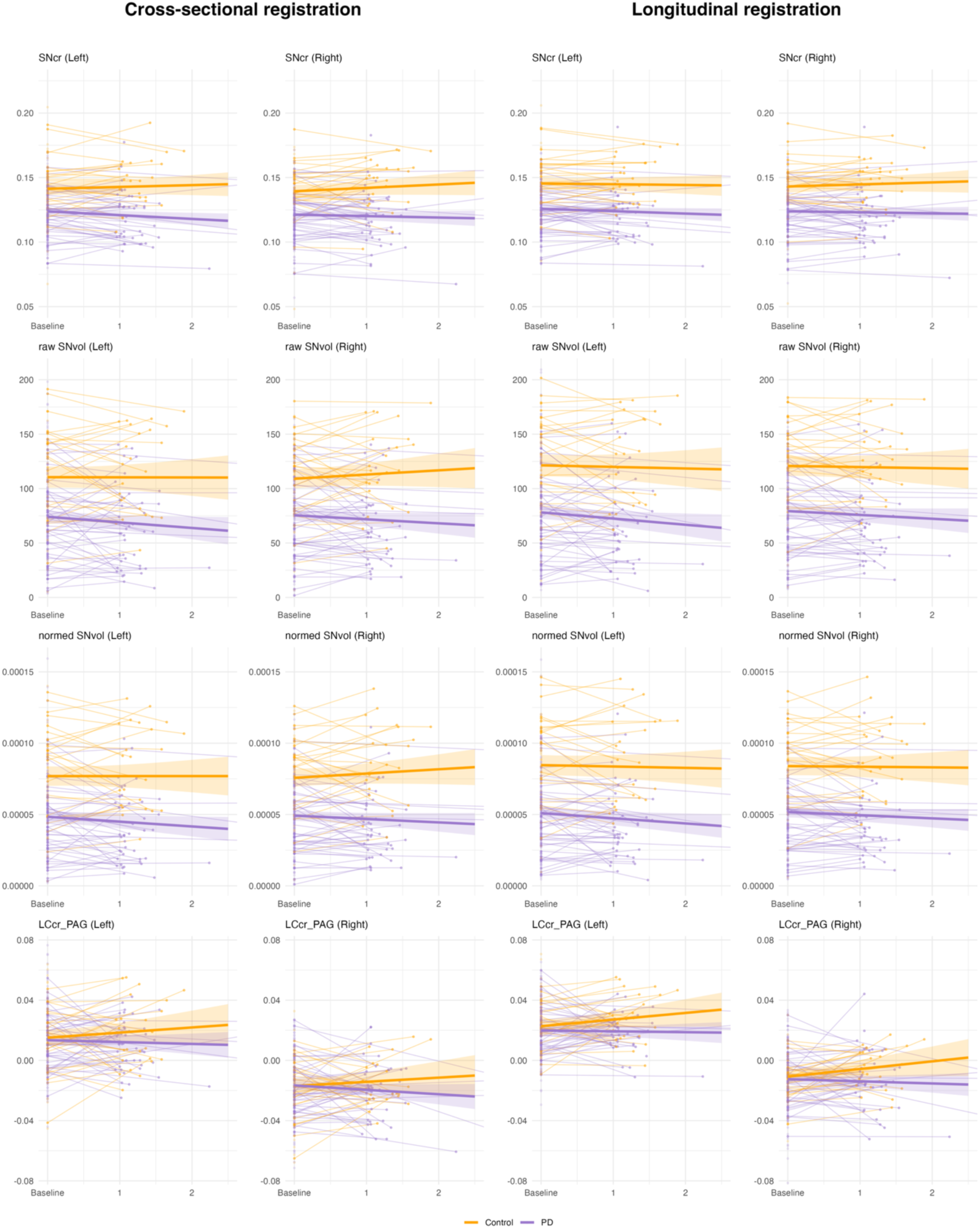
Cross-sectional- (left) and longitudinal-registration-derived (right) NM-MRI metrics for PD (purple) and controls (orange). Individual longitudinal trajectories are shown as spaghetti plots, with baseline-only observations included as points. Solid lines represent linear mixed-effects model predictions and shaded ribbons indicate 95% confidence intervals. Models were fit using all available baseline and longitudinal data. The x-axis is visually clipped to 2.5 years for display purposes.

Figure 8 shows PD-control effect sizes for each NM-MRI metric derived using cross-sectional and longitudinal registration approaches at baseline and follow-up visits, and for annualized change. At baseline and follow-up, both XS and LNG-derived SN metrics exhibited large PD-control effect sizes. LNG-derived SN metrics yielded more negative PD-control effect sizes than XS-derived metrics across all SN measures, with the largest differences observed for raw and normalized SN_vol_ metrics. SN effect sizes were generally larger at follow-up than at baseline for both registration approaches.

**Figure 8:**
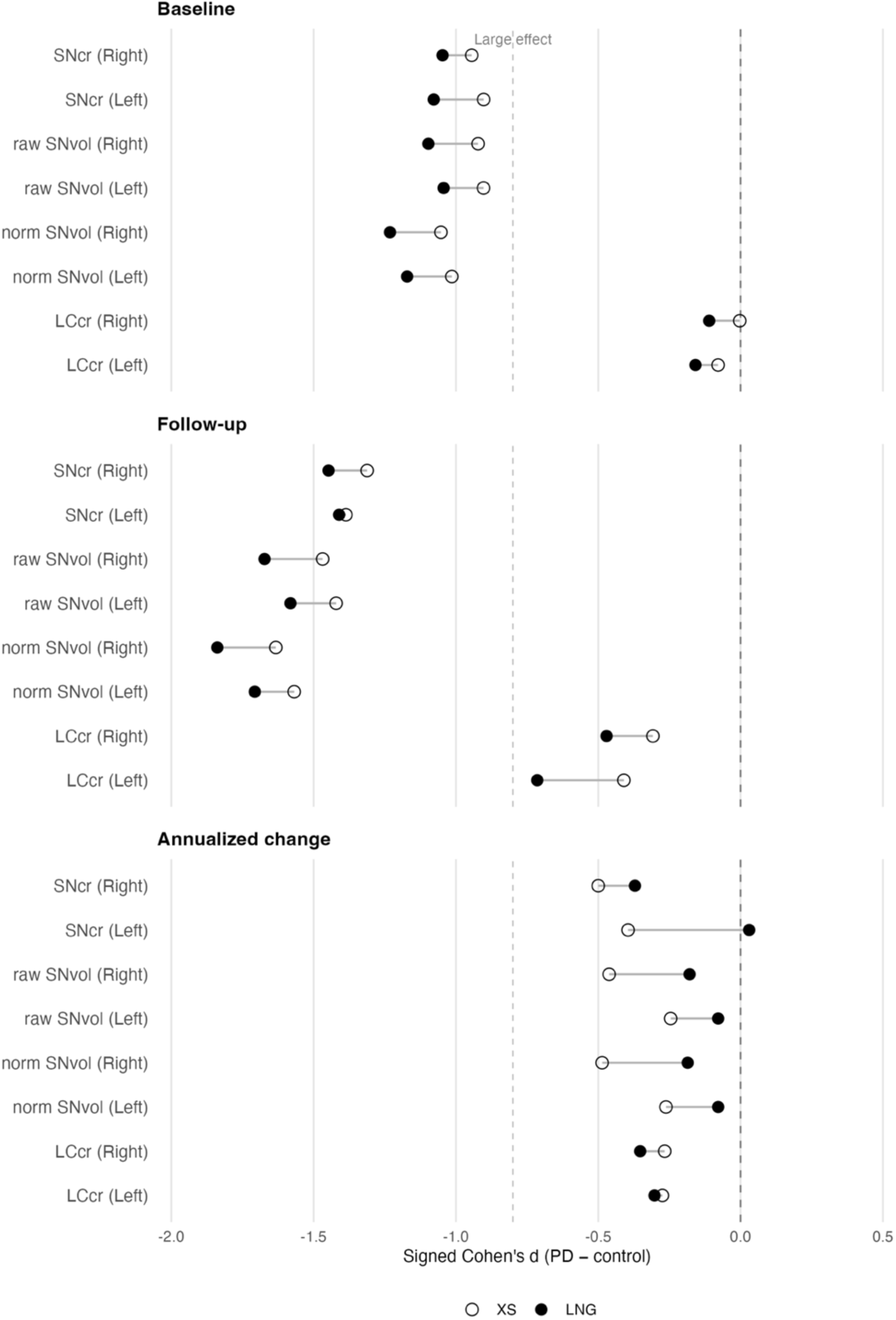
PD-control effect sizes for each NM-MRI metric derived from XS and LNG registration approaches. Signed Cohen’s *d* values (PD–control) are shown for baseline (top), follow-up (middle), and annualized change (bottom). Negative values indicate lower NM-MRI metrics in PD relative to controls. Open circles represent PD-control effect sizes for XS-derived NM-MRI metrics and filled circles represent PD-control effect sizes for LNG-derived NM-MRI metrics. The dashed vertical line at *d* = −0.8 indicates a large effect size.

At baseline, LC_CR_ PD-control effect sizes were small regardless of registration approach (|d| < 0.2). At follow-up, LC_CR_ effect sizes increased for both XS-derived (left: d = −0.41; right: d= −0.31) and LNG-derived metrics (left: d = −0.71; right: d= −0.47), with more negative effect sizes observed for LNG-derived metrics.

For annualized change, PD-control effect sizes for SN metrics were generally more negative for XS-derived metrics, whereas LNG-derived effect sizes were closer to zero. LC_CR_ annualized change effect sizes remained small for both registration approaches.

Figure 9 shows the percent change in residual SD for LNG-derived metrics relative to XS-derived metrics. Residual SD was lower for LNG-derived SN_CR_ (left: −15.1%; right: −11.0%) and LC_CR_ (left: −27.8%; right: −12.3%) metrics, whereas differences between registration approaches were negligible (<1%) for raw and normalized SNvol metrics. The largest reduction in residual SD was observed for left LC_CR_ (−27.8%). This metric which was also the only one showing a significant difference in residual variance between registration approaches (p < 0.001).

**Figure 9:**
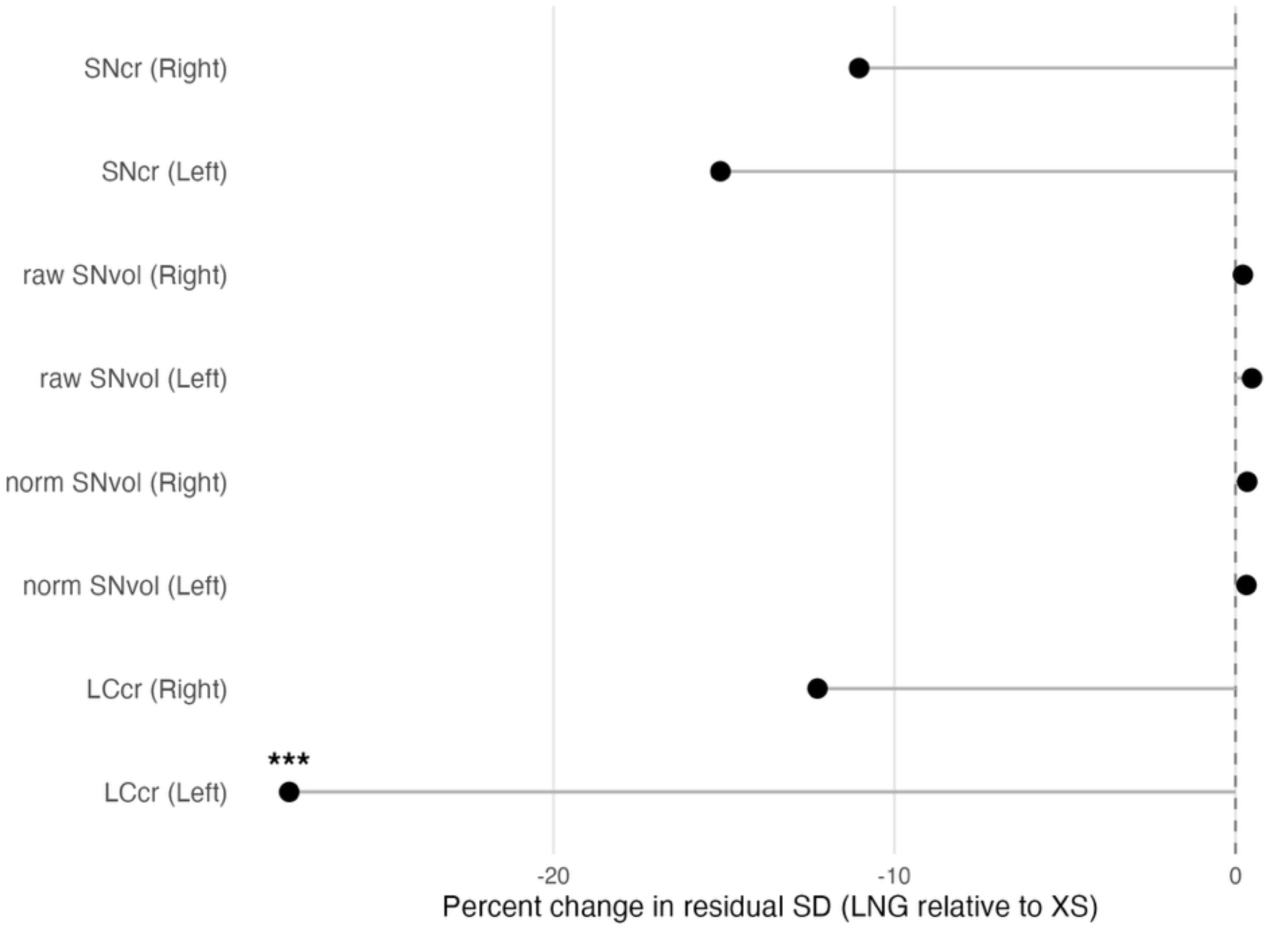
Change in unexplained model variability with LNG registration. Percent change in residual SD is shown for each NM-MRI metric, calculated as LNG relative to XS. Negative values indicate lower residual SD for LNG-derived metrics relative to XS-derived metrics. The dashed vertical line indicates no difference between registration approaches.

## 4.0 Discussion

Overall, the findings indicate that the proposed longitudinal processing framework improves the technical quality of NM-MRI measurements by reducing processing-related variability while preserving the underlying biological interpretation of the data. The greatest benefits were observed in measures of precision, including reductions in within-subject variability and residual variance, with generally stronger PD-control discrimination at each visit following longitudinal registration. However, these technical improvements did not translate into significantly different estimates of longitudinal disease progression over the follow-up interval examined. The following sections discuss the contribution of each processing component to these findings.

### Addressing Slice-wise Intensity Variability in NM-MRI

The lack of visually apparent cross-talk artifacts in the QPN dataset indicates that the interleaved T1w TSE acquisition effectively minimized slice cross-talk under the current protocol. However, qualitative inspection of an external dataset demonstration that slice-by-slice intensity normalization (*inormalize*) substantially reduces striping artifacts when present, supporting its utility as a general preprocessing step.

Quantitatively, *inormalize* produced a small but significant reduction in brainstem voxel intensity standard deviations in control group scans. The largest voxel intensity deviations were observed at superior and inferior slices. Reductions at inferior slices likely reflect the influence of the neck saturation band applied at acquisition. Together, these findings suggest that slice-by-slice normalization reduces slice-dependent intensity variability, particularly at volume boundaries, supporting inclusion in the proposed pipeline.

### Mitigating Systematic Bias for LC NM-MRI

A significant leftward signal asymmetry was observed in the LC, consistent with previous reports with Siemens systems (Trujillo et al., 2024). Similar asymmetries were also observed in non-NM-rich regions (LC-adj, CP) suggesting that the effect is unlikely to reflect true LC lateralization. In contrast, no consistent lateral bias was observed in the SN, although the reason for this remains unclear.

N4 bias field correction did not reduce LC signal asymmetry, and an inspection of the estimated correction fields revealed no meaningful left-right gradients at the level of the LC. Together with observations from independent B_1_ field maps acquired on the same scanner, these findings suggest that the asymmetry is likely driven, at least in part, by scanner-related B_1_-related effects that are not adequately modeled by conventional bias field correction. Forcing a correction on bilateral LC structures that are millimetres apart would require highly localized adjustments that risk distorting adjacent tissue intensities.

Because the asymmetry was present in both PD and controls, it is suggested to express LC NM-MRI metrics relative to the control cohort using z-scores to mitigate this systematic bias by incorporating it into the reference distribution. In addition, the observed rightward CP bias supports the use of pooled CP intensities for SN metric quantification, avoiding potential side-dependent biases in SN NM-MRI metrics.

### Longitudinal Registration Improves Measurement Stability and Precision

The principal finding of this study was that longitudinal registration reduced measurement variability and improved precision of NM-MRI metrics relative to conventional cross-sectional processing. In controls, annualized-change SD was significantly reduced by 25 – 36% for SN_CR_ and by 27 – 34% for LC_CR_PAG_ (Table 3). Because minimal biological change is expected in controls over the follow-up interval, these reductions likely reflect decreased measurement noise rather than attenuation of true longitudinal effects. Similar significant improvements were not observed for volume-based metrics, although variability was consistently lower in LNG-derived metrics.

The residual variance analysis was used as a complementary measure of precision rather than a direct measure of reliability, and has independently supported these findings. As shown in Figure 8, the longitudinal approach reduced residual SD for SN_CR_ and LC_CR_ metrics, with the largest reduction observed for left LC_CR_, whereas residual variability differed minimally between registration approaches for raw and normalized SN volume metrics. Together, the annualized change and residual variance analyses converged on the same conclusion: longitudinal registration reduces technical variability across imaging timepoints and improves the precision of contrast-based NM-MRI measurements.

PAG-based LC normalization also demonstrated greater longitudinal stability than PT-based normalization (Table 3), supporting its use as the preferred reference region for longitudinal LC quantification. Notably, the longitudinal approach improved LC measurement precision despite the persistence of unresolved leftward LC signal asymmetry and the lack of benefit from N4 bias field correction. This suggests that longitudinal registration addresses an important source of measurement variability that is independent of scanner-related signal asymmetry.

Nevertheless, the increasing LC trajectories observed in both groups (Figure 7), together with the persistence of unresolved signal asymmetries, indicate that the longitudinal LC changes should be interpreted cautiously. While LC NM-MRI metrics derived from this cohort have demonstrated clinically meaningful associations (Sun et al., 2025), cross-sectional analyses relative to a control reference distribution may currently be easier to interpret biologically than longitudinal LC changes at 3T.

### Differential Effects of Registration Approach on Contrast- and Volume- Based Metrics

Longitudinal registration affected contrast- and volume-based NM-MRI metrics differently. Although the longitudinal approach generally produced larger PD-control effect sizes compared to the cross-sectional approach at baseline and follow-up (Figure 8), the largest improvements were observed for raw and normalized SN volume metrics, consistent with the significant *method × group* interactions identified in the mixed-effects models (Table 4). In contrast, contrast-based LNG-derived metrics showed comparatively modest changes in PD-control discrimination compared to XS-derived metrics despite exhibiting the greatest reductions in annualized change variability (Table 3).

An apparent paradox of the current findings is that the largest improvements in PD-control discrimination were observed for SN volume metrics, despite the strongest stability gains occurring for contrast-based metrics. One possible explanation is that volume metrics are particularly sensitive to small differences in ROI boundary placement, given that the metric relies on a CP intensity-based threshold. In contrast, contrast-based metrics are calculated using median intensity values, which are more robust against minute registration differences compared with intensity average- or percentile-based approaches (Madge et al., 2023a). Consequently, small improvements in registration precision may disproportionately affect SN volume estimates, as voxels near the threshold can be reclassified as either NM-positive or NM-negative. This may explain why the largest gains in PD-control discrimination were observed for volume-based metrics despite more modest improvements in measurement stability.

### Improved Precision Did Not Translate into Improved Longitudinal Sensitivity

Despite reducing variability, longitudinal registration did not significantly improve longitudinal PD-control group separation relative to the cross-sectional approach. Significant *group × time* interactions were observed for bilateral SN_CR_ and LC_CR_ metrics under the reference cross-sectional approach (Table 4), whereas significant *method × group × time* interactions were not observed for any NM-MRI metric. Consistent with these findings, the fitted trajectories derived from the mixed-effects models did not demonstrate substantial differences in longitudinal group separation between registration approaches (Figure 7).

One possible explanation for the absence of significant *method × group × time* interactions is the availability of only two imaging timepoints per participant. With two visits, each subject’s trajectory is constrained to a single linear difference between baseline and follow-up, limiting the ability to characterize nonlinear change, plateauing, or consistency of change over time. In contrast, three or more visits would provide additional information about the shape and reliability of within-subject trajectories. Moreover, the *method × group × time* interaction represents a higher-order test of whether registration approach modifies longitudinal PD-control differences, which generally requires larger samples and longer follow-up intervals than tests of within-visit group separation.

Beyond considerations of statistical power, longitudinal NM-related changes may be intrinsically difficult to detect in established PD. The large cross-sectional PD-control differences observed across SN metrics (Figure 8), together with previous reports of substantial NM loss by onset of motor symptoms (Zecca et al., 2001), suggest that much of the detectable NM signal loss may occur during prodromal or early disease stages, leaving comparatively smaller changes to be detected following diagnosis and during the relatively short follow-up interval examined here with PD participants.

Interestingly, the registration approach associated with larger cross-sectional PD-control effect sizes also produced lower annualized change variability (Table 3) and smaller annualized change effect sizes (Figure 8). Conversely, several SN metrics exhibited larger annualized change effect sizes under the cross-sectional approach despite the greater longitudinal variability. Together, these findings suggest that some apparent longitudinal effects may be influenced by measurement instability rather than biological change, and that longitudinal registration may reduce noise-driven apparent change while preserving disease-related group differences.

Importantly, reduced technical variability remains valuable even when it does not improve longitudinal group separation over a short follow-up interval. Improved measurement precision may facilitate detection of subtler biological effects and biomarker relationships in future studies. As NM-MRI is increasingly being explored as a progression biomarker and clinical trial endpoint, reducing processing-related variability may be particularly important for future longitudinal studies.

### Limitations and Future Work

Several limitations should be considered. First, all analyses were performed using data acquired on a single scanner using a single NM-MRI acquisition protocol, and the generalizability of these findings to other scanners, field strengths, and NM-MRI sequences remains to be established. Second, the longitudinal cohort contained only two imaging timepoints per participant, limiting the ability to characterize nonlinear trajectories and reducing sensitivity to subtle longitudinal effects. Third, the median follow-up interval of approximately one year may be insufficient to capture substantial NM-related changes in established PD. The longitudinal subset was also modest in size, sex-imbalanced, and follow-up intervals were variable due to COVID-19-related disruptions, reducing power to detect small annualized changes, sex differences, and higher-order interaction effects.

Persistent LC signal asymmetry could not be physically corrected and instead it was suggested to mitigate this bias using control-referenced normalization. In addition, because analyses were performed on unlateralized data, left-right differences should not be interpreted as evidence of biological laterality.

Future work should evaluate the proposed pipeline in larger multi-site cohorts with additional longitudinal timepoints and longer follow-up intervals. Higher-resolution LC imaging and direct B₁ field characterization may help further address unresolved sources of LC measurement variability.

## 5.0 Conclusion

This study presents and validates a longitudinal NM-MRI processing framework designed to reduce processing variability and improve the robustness of longitudinal NM-MRI measurements in the SN and LC.

Validation experiments identified several processing choices that improved measurement stability and consistency. Slice-wise intensity normalization reduced slice-dependent intensity variation, systematic LC signal asymmetry was characterized, and PAG-based normalization provided more stable longitudinal LC measurements than PT-based normalization. Longitudinal registration improved the stability and precision of NM-MRI measurements, particularly for contrast-based SN and LC metrics, as demonstrated by reduced annualized-change variability in controls, and reduced residual variability in longitudinal PD-control comparisons.

Although longitudinal registration did not significantly improve detection of longitudinal PD-control differences, it consistently strengthened PD-control separation at individual visits while reducing apparent longitudinal variability. Together, these findings suggest that the proposed framework is effective at reducing processing- and registration-related variability that may otherwise be misinterpreted as biological change.

Overall, the results support the use of longitudinal registration and the proposed processing framework for longitudinal NM-MRI studies of PD. By improving measurement stability and reducing technical sources of variability, the framework provides a more robust foundation for investigating NM-related neurodegeneration in the SN and LC over time.

## Data Availability

The longitudinal NM-MRI processing pipeline developed in this study is openly available at https://github.com/NIST-MNI/nm-longitudinal-pipeline.

The PD126 template is publicly available at https://nist.mni.mcgill.ca/multi-contrast-pd126-and-ctrl17-templates/.

Baseline imaging data are publicly available at https://doi.org/10.5281/zenodo.17246063.

Longitudinal imaging, associated clinical data, and anatomical labels are not publicly available at the time of publication but may be made available to qualified researchers upon reasonable request and approval by the Quebec Parkinson Network.

## Acknowledgements

This work acknowledges the Quebec Parkinson Network team for their essential role in coordinating longitudinal imaging and clinical data collection.

We also acknowledge funding from *Weston Family Foundation* Rapid Response Grant RR171117 and the *Famille Louise et Andre Charron*.

